# Statistical power in clinical trials of interventions for mood, anxiety, and psychotic disorders

**DOI:** 10.1101/2021.10.12.21264893

**Authors:** Ymkje Anna de Vries, Robert A. Schoevers, Julian P. T. Higgins, Marcus R. Munafò, Jojanneke A. Bastiaansen

## Abstract

**Background:** Previous research has suggested that statistical power is suboptimal in many biomedical disciplines, but it is unclear whether power is better in trials for particular interventions, disorders, or outcome types. We therefore performed a detailed examination of power in trials of psychotherapy, pharmacotherapy, and complementary and alternative medicine (CAM) for mood, anxiety, and psychotic disorders.

**Methods:** We extracted data from the Cochrane Database of Systematic Reviews (Mental Health). We focused on continuous efficacy outcomes and estimated power to detect standardized effect sizes (SMD=0.20-0.80, primary effect size SMD=0.40) and the meta-analytic effect size (ES_MA_). We performed meta-regression to estimate the influence of including underpowered studies in meta-analyses.

**Results:** We included 216 reviews with 8809 meta-analyses and 36540 studies. Statistical power for continuous efficacy outcomes was very low across intervention and disorder types (overall median [IQR] power for SMD=0.40: 0.33 [0.19-0.54]; for ES_MA_: 0.15 [0.07-0.44]), only reaching conventionally acceptable levels (80%) for SMD=0.80. Median power to detect the ES_MA_ was higher in TAU/waitlist-controlled (0.54-0.66) or placebo-controlled (0.15-0.40) trials than in trials comparing active treatments (0.07-0.10). Meta-regression indicated that adequately-powered studies produced smaller effect sizes than underpowered studies (B=-0.06, p=0.008).

**Conclusions:** Power to detect both fixed and meta-analytic effect sizes in clinical trials in psychiatry was low across all interventions and disorders examined. As underpowered studies produced larger effect sizes than adequately-powered studies, these results confirm the need to increase sample sizes and to reduce reporting bias against studies reporting null results to improve the reliability of the published literature.

## Introduction

Mental disorders such as depression, anxiety disorders, and psychosis are responsible for a large proportion of the global disease burden (Whiteford *et al*., 2013). Effective treatment options are, however, available -- mainly various forms of pharmacotherapy and psychotherapy (Huhn *et al*., 2014), although some complementary and alternative medicine (CAM) treatments (e.g., mindfulness) also appear to be effective for some disorders (Kuyken *et al*., 2016; Asher *et al*., 2017). Consistent with the ideals of evidence-based medicine (EBM), treatment efficacy is supported by randomized controlled trials (RCTs), the gold standard for high-quality evidence. However, there has been increasing concern that the evidence base that EBM depends on is distorted. The efficacy of antidepressants and antipsychotics, for instance, has been inflated by reporting bias (Turner *et al*., 2008, 2012; Roest *et al*., 2015; de Vries *et al*., 2018), and the same is probably true for psychotherapy (Driessen *et al*., 2015; de Vries *et al*., 2018). Problems in trial design can also lead to stacking the deck in favor of a treatment (Heres *et al*., 2006; Leichsenring *et al*., 2016) or to difficulty generalizing results to clinical practice (Lorenzo-Luaces *et al*., 2018). Here, we focus on one particular problem in trial design, namely inadequate statistical power.

Statistical power is the probability of detecting an effect of a specific size if that effect is actually present. The threshold for adequate power is conventionally set at 80% (Cohen, 1992). Inadequate statistical power increases the likelihood of falsely concluding that an intervention is not effective as well as the likelihood that statistically significant effects represent false positive findings. The problem of low power in individual trials can in principle be resolved through meta-analysis: by combining underpowered studies, a well-powered meta-analysis can result in a precise estimate (Guyatt *et al*., 2008). However, reporting bias is ubiquitous (Song *et al*., 2010), and the problem of low power is more pernicious when combined with reporting bias. While underpowered studies are as likely to yield an *underestimate* of the true effect size as they are to yield an *overestimate*, reporting bias filters out (statistically non-significant) underestimates. This may result in a literature dominated by false-positives and inflated effect sizes.

Low power to detect relevant effect sizes has previously been demonstrated for studies in neuroscience (Button *et al*., 2013), biomedicine (Dumas-Mallet *et al*., 2017), and the social sciences (Smaldino and McElreath, 2016). An examination of the Cochrane Database of Systematic Reviews (CDSR) by Turner and colleagues found that median power to detect a relative risk reduction of 30% was only 14% in trials specifically for mental health (comparable with 13% for medicine in general). Furthermore, effect sizes were reduced by 12-15% when only adequately-powered studies were considered (Turner *et al*., 2013). On the other hand, a study of psychotherapy trials for depression reported that average power to detect the meta-analytic effect size was somewhat better, at 49% (Flint *et al*., 2015).

These findings might indicate that there are large differences in median power depending on the intervention type, but there are other potential explanations. For instance, Turner and colleagues only included binary outcomes, even though the primary outcome in psychiatric trials is usually continuous (e.g., decrease in symptoms). Examining only binary outcomes, for which trials were not powered, could result in a lower estimate of power than for continuous outcomes. Furthermore, although Turner and colleagues examined both power to detect a fixed effect size and power to detect the meta-analysis-specific effect size, the latter was only examined across all trials, regardless of medical specialty or intervention type. This may be important because effect sizes vary widely. Comparing antidepressants with placebo, for instance, the standardized mean difference (SMD) is around 0.3 (Turner *et al*., 2008; Roest *et al*., 2015), while the SMD for psychotherapy compared to waitlist for depression is around 0.9 (Cuijpers *et al*., 2010). However, the SMD of psychotherapy compared to more active control conditions (e.g. treatment as usual) is much lower and similar to that for antidepressants vs. placebo (Cuijpers *et al*., 2010). As statistical power primarily depends on sample size and effect size, calculating power based on the same effect size across disorders, interventions, and comparators could lead to either an underestimate or overestimate of power for interventions that are actually markedly more or less effective than the chosen effect size. While the “true” effectiveness of an intervention cannot be known, meta-analytic effect sizes can be used as a (noisy) proxy. The above-mentioned studies, for instance, suggest that power might be somewhat better in psychotherapy trials, but this may be due to the comparators used. A comparison of power in trials for different intervention types within the domain of psychiatry has not yet been done.

In this study, therefore, we performed a detailed examination of statistical power to detect both fixed and meta-analysis-specific effect sizes in trials of psychotherapy, pharmacotherapy, and CAM for mood, anxiety, and psychotic disorders. We focused on continuous efficacy outcomes, but also examined other outcomes (binary efficacy and safety). We also examined whether statistical power is increasing over time, as previous studies have suggested that the problem of low power is either not improving at all or improving only slightly (Smaldino and McElreath, 2016; Lamberink *et al*., 2018). Finally, we examined whether inclusion of underpowered studies in meta-analyses affects estimated effect sizes, which offers some insight into whether low power is associated with inflated effect sizes. This fine-grained comparison of statistical power can provide clinicians and researchers with a better sense of where the problem of low power is most acute and hence with starting points for improvements.

## Methods

### Preregistration

This study was preregistered after we received the data, but before performing any analyses (osf.io/hgaec).

### Data source and selection

We obtained permission from the Cochrane Collaboration to use Cochrane data for this study. We received an export of currently published systematic reviews of interventions in the Mental Health area in RevMan (RM5) format in October 2017. We extracted the following information from each review file: review title, comparison, outcome, subgroup, names of group 1 and group 2, study names, type of effect measure (e.g., SMD), effect size with confidence interval and standard error (if available), number of events in group 1 and group 2 (for binary outcomes), sample size in group 1 and group 2. Each combination of comparison, outcome, and subgroup made up a single meta-analysis within a review.

Reviews were categorized by topic and intervention by one author (YV, checked by JB). We categorized each review into one of three categories: mood disorders, anxiety disorders, and psychotic disorders. Reviews that did not fit one of these categories (e.g., interventions for aggression) or fit multiple categories (e.g., depression and anxiety) were excluded; however, if the review contained meta-analyses that focused on one specific category, we assigned individual meta-analyses to the applicable category. We also assigned each review to one of three categories of treatment: pharmacotherapy (PHT), psychotherapy (PST), or CAM (defined based on a topic list provided for the Cochrane Collaboration (Wieland *et al*., 2005)). Reviews that did not clearly fit one of these categories were excluded. Reviews or meta-analyses that investigated combination PHT and PST were assigned to PST if the comparator was PHT, to PHT if the comparator was PST, or excluded if the comparator was treatment as usual.

We excluded meta-analyses that only included a single study and meta-analyses that were not analyzable because the event rate was 0. We also excluded meta-analyses that compared the experimental intervention to unusual control interventions (i.e., that did not match pharmacotherapy, psychotherapy, CAM, placebo, treatment as usual, waitlist, or a combination of these). Meta-analyses were assigned to one of four categories: (1) continuous efficacy outcome (e.g., symptom questionnaires), (2) binary efficacy outcome (e.g., relapse), (3) continuous safety outcome (e.g., weight gain), or (4) binary safety outcome (e.g., occurrence of nausea). Efficacy vs. safety was determined by one author (YV) based on the description of the outcome (with any unclear outcomes checked by JB), while binary vs. continuous was determined based on the effect measure (odds ratio [OR]/risk ratio/risk difference vs. (standardized) mean difference). We chose the continuous efficacy measure as our primary outcome, as this is commonly used as primary outcome in psychiatry.

### Effect size and power calculations

We first re-calculated meta-analyses using a mean difference, risk difference, or risk ratio as an outcome to use the SMD or OR instead. We used mean differences, standard errors, and sample sizes to calculate SMDs and event rates and sample sizes to calculate ORs. Random-effects meta-analysis was performed using restricted maximum likelihood estimation (REML) via the *rma* command from the *metafor* package (2.0-0) in R (3.5.0). Most effect sizes were negative (for continuous outcomes) or smaller than 1 (for binary outcomes). We multiplied all effect sizes for continuous outcomes by −1 and took the inverse of all ORs, so that effect sizes are usually positive (for continuous outcomes) or greater than 1 (for binary outcomes). For active vs. active comparisons (e.g., antidepressant vs. another antidepressant), we used the absolute effect size or the inverse of the OR (if OR<1), as experimental and comparator conditions can be seen as interchangeable.

We estimated the power of each study to detect small to large effect sizes (SMD=0.20, 0.40, 0.60 or 0.80, or the roughly equivalent OR=1.5, 2.0, 3.0, and 4.5, using the formula log(OR)=SMD x π/3 and rounded to the nearest 0.5). We set SMD=0.40 as our primary effect size, as this is close to the mean effect size for psychiatric treatments in general (Leucht *et al*., 2012; Huhn *et al*., 2014). We also estimated each study’s power to detect the effect size of the meta-analysis it was included in (ES_MA_). We calculated the power for each study using the *pwr*.*t2n*.*test* command for continuous outcomes and the *pwr*.*2p2n*.*test* command for binary outcomes (*pwr* package (1.2-2) in R). To examine trends in power to detect SMD=0.40 over time, we plotted median power against publication year.

### Meta-regression analysis of adequate power

Following Turner and colleagues (Turner *et al*., 2013), we investigated the impact of underpowered studies on the estimated effect size of continuous efficacy outcomes. We selected meta-analyses that included ≥5 studies, of which ≥2 were adequately powered and ≥1 was underpowered. We defined adequate power as being ≥80%, provided that ≥100 meta-analyses qualified for inclusion using this cut-off, and as ≥50% if not. For each group of studies in a meta-analysis, we fit a random-effects meta-regression model (using *rma*) with a term for “adequate power”. Subsequently, we used random-effects meta-analysis to summarize the effect of adequate power across meta-analyses.

### Sensitivity analyses

We performed several planned sensitivity analyses for the continuous efficacy outcome. First, we calculated power to detect the ES_avg_, rather than the ES_MA_. We defined the ES_avg_ as the meta-analytic average effect size of all meta-analyses for each combination of outcome (efficacy vs. safety), outcome type (binary vs. continuous), experimental group, and comparator group. Although the ES_MA_ is a proxy for the ‘true’ effect size of a specific intervention for a specific outcome (e.g., paroxetine vs. placebo for depressive symptoms), it may be too noisy if meta-analyses only include a few studies. The ES_avg_, on the other hand, is less specific because it is aggregated across similar interventions and outcomes (e.g., any pharmacotherapy vs. placebo for any continuous efficacy outcome), but also more stable. Second, we recalculated power to detect the ES_MA_ after excluding meta-analyses with very small effect sizes (ES_MA_<0.2). Third, because the ES_MA_ might be inflated due to publication bias, we recalculated power using the effect size of the largest trial in each meta-analysis. Finally, because studies could be included in multiple meta-analyses, we recalculated power for the standard effect sizes and the ES_MA_, while only including each study once.

## Results

### Data selection and characteristics of included reviews

Figure 1 shows a flow chart of the selection process. We received 349 reviews, of which 315 included usable data. After exclusion of ineligible reviews and meta-analyses, we retained 216 reviews with 8809 meta-analyses. Among these meta-analyses, 1993 concerned continuous efficacy outcomes (primary outcome), 268 continuous safety outcomes, 1912 binary efficacy outcomes, and 4636 binary safety outcomes. The final dataset contained 36540 observations (i.e., studies), but many studies were included in multiple meta-analyses; there were only approximately 3888 distinct studies. Each review included on average 40.8 meta-analyses (median=23, range=1-436), while each meta-analysis included on average 4.1 studies (median=3, range=2-80).

**Figure 1:**
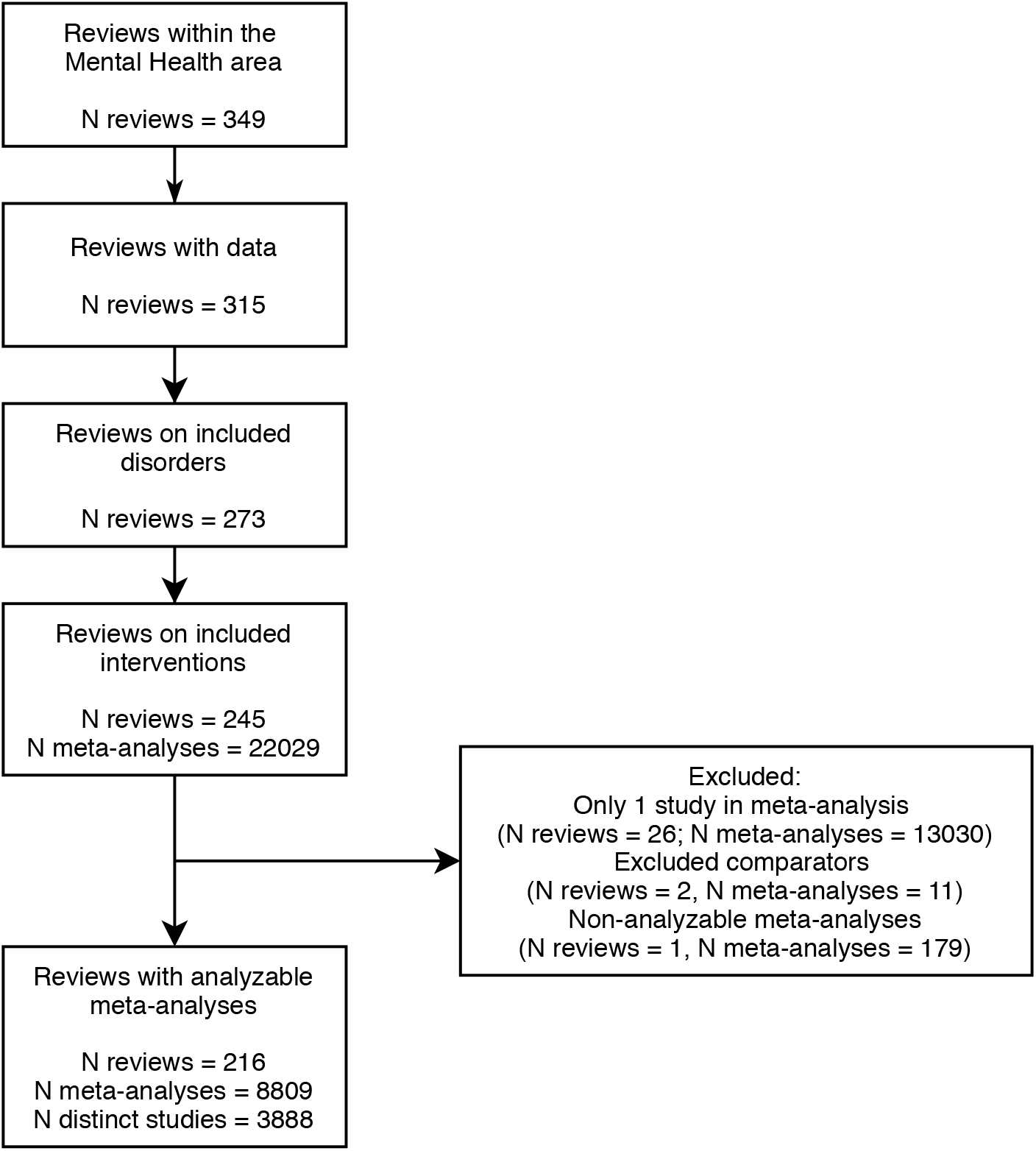
Flow chart of the selection process

### Effect sizes and power for continuous efficacy outcomes

Figure 2 shows the distribution of ESMA for continuous efficacy outcomes, by disorder and intervention category (see Supplemental Table 1 for detailed information). The overall median effect size was 0.24 (interquartile range [IQR] = 0.09-0.46). Meta-analyses for anxiety disorders had larger effect sizes (median [IQR] = 0.35 [0.15-0.58]) than those for mood disorders (median [IQR] = 0.19 [0.07-0.39]) or psychotic disorders (median [IQR] = 0.17 [0.08-0.37]). Meta-analyses of CAM interventions also had larger effect sizes (median [IQR] = 0.47 [0.19-0.69]) than meta-analyses of PHT (median [IQR] = 0.21 [0.08-0.41]) or PST (median [IQR] = 0.28 [0.11-0.53]). These differences may be related, at least in part, to the comparators frequently used. Only 19% of meta-analyses for anxiety disorders compared the intervention with another similarly active comparator, compared to 45% of those for mood disorders and 76% of those for schizophrenia. Similarly, only 19% of CAM meta-analyses compared the intervention with another similarly active comparator, compared to 51% of PHT meta-analyses and 42% of PST meta-analyses. Effect sizes were larger for comparisons of active therapy with TAU/waitlist (median ES_MA_=0.58-0.63) or placebo (median ES_MA_=0.38-0.41), and of combination therapy with monotherapy (median ES_MA_=0.28-0.54), than for comparisons of monotherapy vs. another monotherapy (median ES_MA_=0.13-0.23) (see Supplemental Table 2).

**Figure 2:**
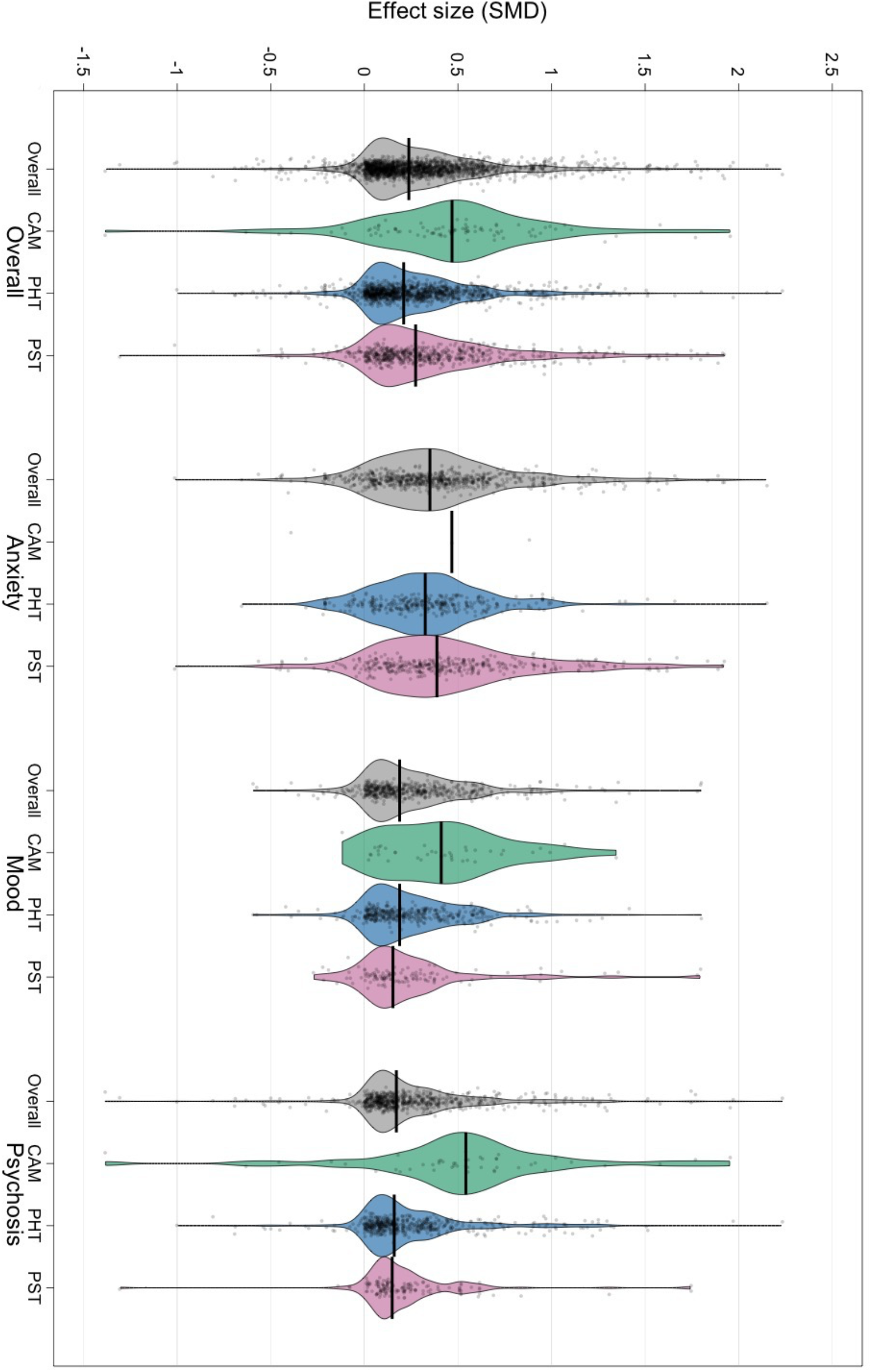
Distribution of standardized effect sizes, by disorder and intervention category

Figure 3 shows the distribution of estimated power to detect SMD=0.40 among studies with continuous efficacy outcomes. Overall, median power was 0.33 (IQR = 0.19-0.54). Median power was slightly higher in studies of PHT (0.34 [0.20-0.59]) than in studies of CAM (0.26 [0.15-0.40]) or PST (0.28 [0.18-0.46]). It was also higher in studies of mood disorders (0.36 [0.20-0.68]) and psychotic disorders (0.36 [0.26-51]) than in studies of anxiety disorders (0.23 [0.17-0.39]) (see Supplemental Table 3 for detailed information). Median power only exceeded the recommended threshold of 80% for SMD=0.80 (0.86 [0.58-0.98]). Consistent with the low median meta-analytic effect size (SMD=0.24), power to detect the ES_MA_ was generally lower than the estimated power to detect an SMD=0.40. Overall power to detect the ES_MA_ was only 0.15 [0.07-0.44]. Consistent with the differences in effect sizes, power to detect the ES_MA_ was generally better in trials using TAU/waitlist (0.54-0.66) or placebo (0.15-0.40) as a comparator than in trials with active vs. active comparisons (0.07-0.10) (see Supplemental Table 4).

**Figure 3:**
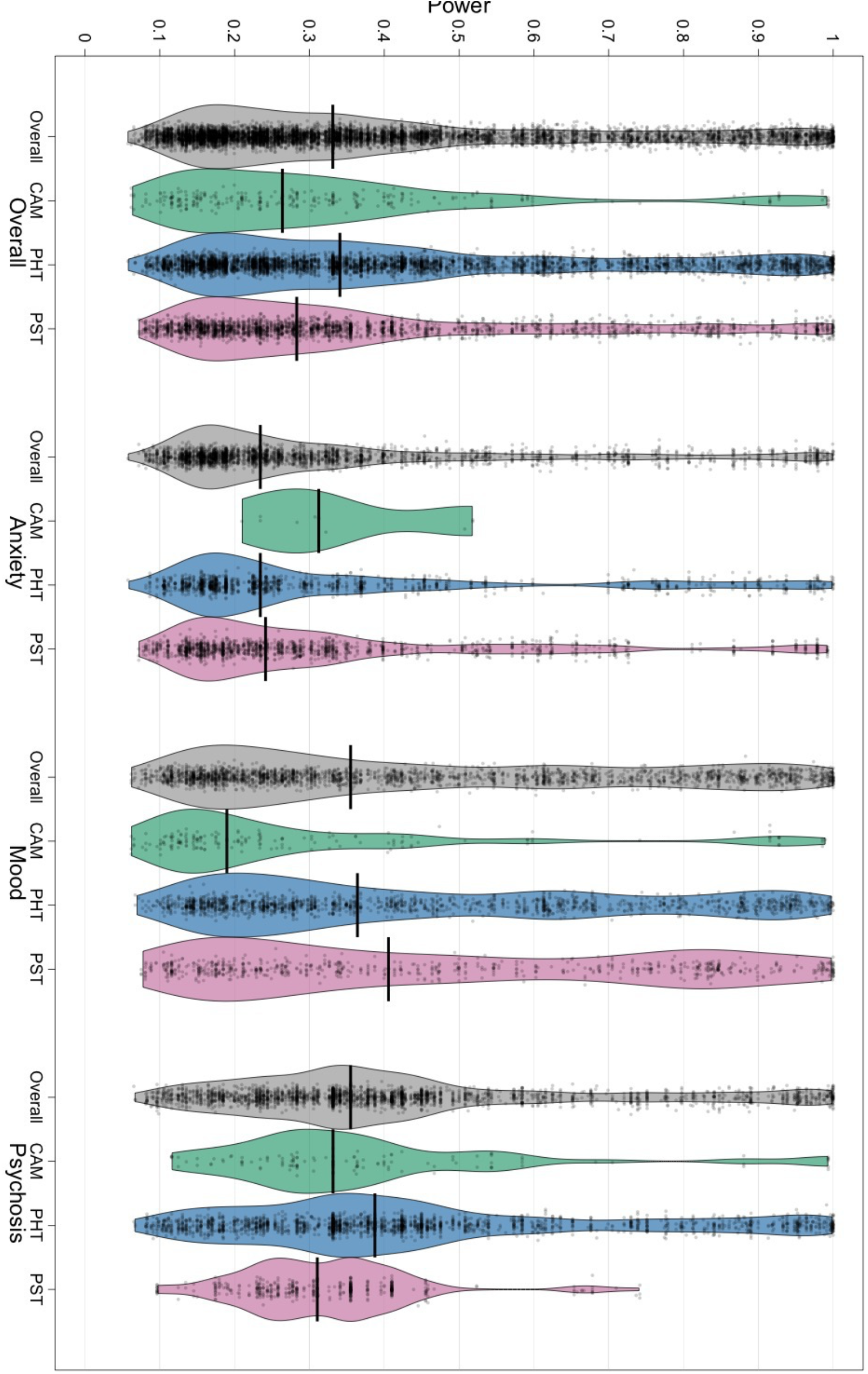
Distribution of power, by disorder and intervention category

Examining the trend in median power to detect an SMD=0.40 over time suggested an increase in power, from a median of around 0.25 from 1960 until 1990, increasing to around 0.40 in recent years (Figure 4). This trend appeared to be present for each intervention type (Supplemental Figure 1).

**Figure 4:**
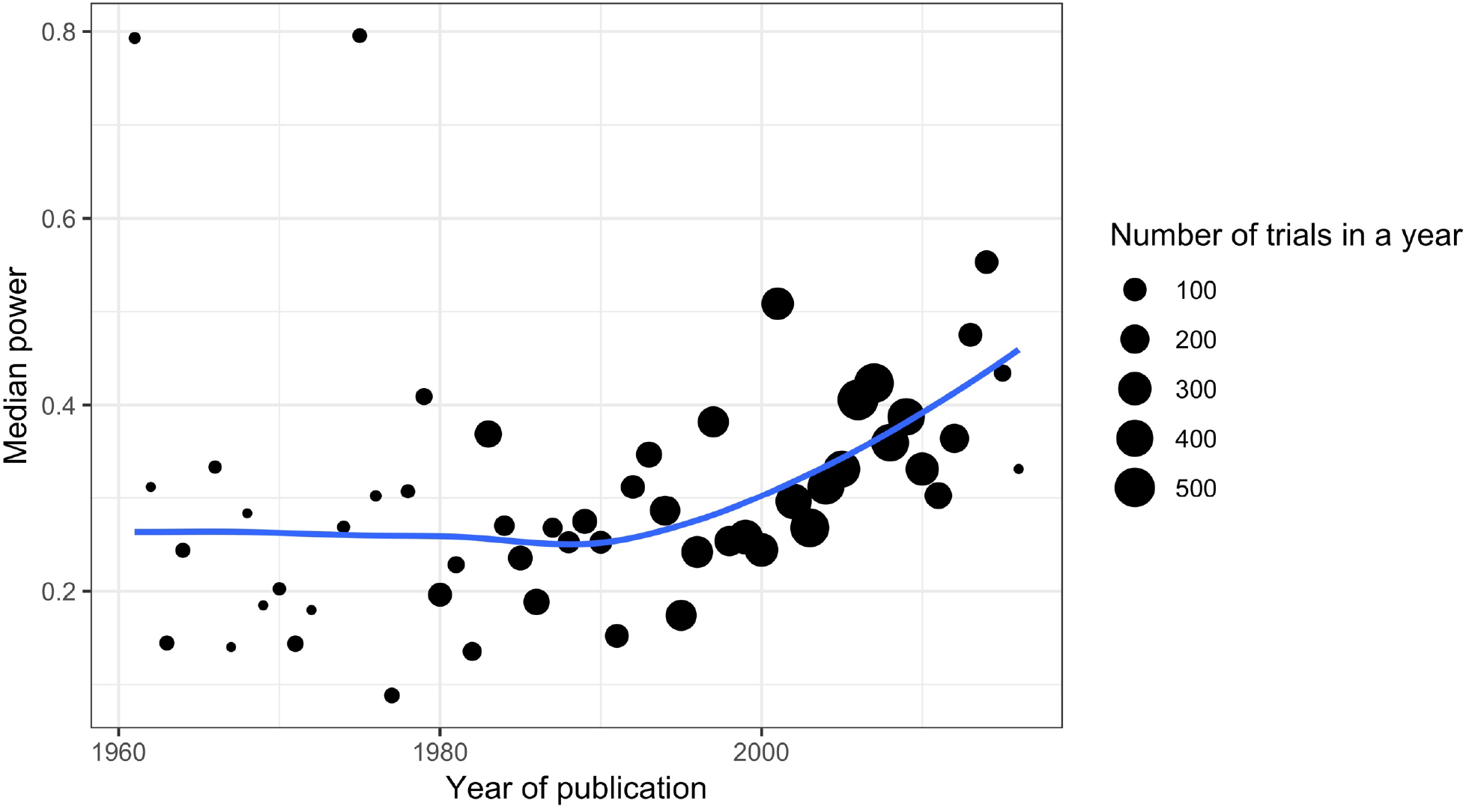
Median power by year of publication (with Loess smoothed line)

### Effect sizes and power for other outcomes

Supplemental Tables 5 through 7 contain the median ES_MA_ by disorder and intervention type for continuous safety outcomes and for binary safety and efficacy outcomes. Supplemental Tables 8 through 10 additionally contain the median ES_MA_ by intervention-comparator combination. Overall, the median ES_MA_ for continuous safety outcomes was SMD=0.13 [IQR=0.03-0.34]. The median ES_MA_ for binary efficacy and safety outcomes was OR=1.27 [0.96-1.99] and OR=1.36 [1.07-1.97], respectively.

Supplemental Tables 11 through 16 provide detailed information on power to detect the full range of effect sizes by disorder and intervention type and by intervention-comparator combination. In brief, median power to detect an SMD=0.40 among trials examining a continuous safety outcome was quite high, at 0.79 [0.39-0.95]. However, median power to detect OR=2.0 was 0.22 [0.15-0.40] for binary efficacy outcomes and 0.21 [0.13-0.38] for binary safety outcomes. Consistent with the low median ES_MA_ for all outcomes, power to detect the ES_MA_ was lower than the power to detect SMD=0.40 or OR=2.0. There were no consistent patterns in power to detect the ES_MA_ based on intervention-comparator combination for safety outcomes. However, for binary efficacy outcomes, patterns mirrored those for continuous efficacy outcomes, with higher power in trials using placebo or TAU/waitlist (0.08-0.51) than in trials with active vs. active comparisons (0.07-0.15).

### Impact of underpowered studies on meta-analyses

Exactly 100 meta-analyses met inclusion criteria at an adequate power cut-off of ≥80%. On average, underpowered studies had an effect size of 0.24, and there was a significant difference in effect size between adequately-powered and underpowered studies (B=-0.06, p=0.008, τ2=0.02, I^2^=49%), indicating that adequately-powered studies resulted in significantly lower effect sizes.

### Sensitivity analyses

We performed several sensitivity analyses for the continuous efficacy outcome (Supplemental Tables 17, 18, and 19). Overall, the ES_avg_ was similar to the median ES_MA_ for most intervention-comparator combinations (Supplemental Table 17), and power to detect the ES_avg_ was also similar to the power to detect the ES_MA_ found in our main analyses, but with less variation (0.14 [0.09-0.25] compared to 0.15 [0.07-0.44]). Exclusion of meta-analyses with very small effect sizes resulted in an increase in overall median power to detect the ES_MA_ (to 0.36 [0.18-0.71], but power remained quite low. Basing the ES_MA_ on the largest trial in a meta-analysis did not meaningfully alter overall median power to detect the ES_MA_ (estimated at 0.13 [0.06-0.40]). Finally, only including each study once reduced the sample size by around 75%, but estimates of power were nearly identical (e.g., overall power to detect SMD=0.40 was 0.33 [0.21-0.59], compared to 0.33 [0.19-0.54] in our main analyses). This suggests that our main analyses were not overly influenced by a small subset of studies included in many meta-analyses.

## Discussion

### Principal findings

In this study, we provide a detailed examination of statistical power in clinical trials in psychiatry. We find that low power is a widespread problem, with relatively small differences among the different disorders and different intervention types. Overall, median power to detect a medium effect size (SMD=0.40) for a continuous efficacy outcome was 0.33, well below recommended levels (80%). Median power to detect the meta-analysis-specific effect size (ES_MA_) was even lower, at only 0.15. There was a clear relationship between the intervention-comparator combination and power to detect the ES_MA_. Trials that compared an active treatment to a less active treatment (e.g., pharmacotherapy vs. placebo or psychotherapy compared to TAU/waitlist) had higher median power to detect the ES_MA_ (0.15-0.66) than trials that compared similarly active treatments (0.08-0.10).

We also examined binary efficacy outcomes as well as binary and continuous safety outcomes. Surprisingly, we found that median power to detect SMD=0.40 was relatively high for continuous safety outcomes (median power=0.79). However, such outcomes (e.g., weight change) were uncommon and almost exclusively used in trials comparing two antipsychotics. It is unclear why median power was relatively high for these outcomes. However, mental health trials are seldom powered specifically to detect safety events; hence, it is more likely that this was a side effect of a selection process, with these outcomes only being included in trials that happened to be large, than a deliberate attempt to adequately power these specific outcomes. In contrast, median power to detect OR=2.0 for binary outcomes was very low, at 0.21-0.22. This is consistent with the fact that larger sample sizes are required to detect a similar effect size for binary outcomes than for continuous outcomes, although we also note that SMD=0.40 and OR=2.0 are only approximately equivalent. Given this, avoiding unnecessary dichotomization of continuous variables (e.g. into remission vs. non-remission) is one way to increase statistical power in clinical trials.

### Implications and comparison with previous literature

It is generally recommended that trials should have a power of 80% to detect a desired effect size. This effect size might be the expected effect size based on previous literature (but note that this is fraught with difficulties (Anderson *et al*., 2017)) or the minimal clinically relevant effect size. Our findings suggest that trialists in the mental health field implicitly work under the assumption that SMD=0.80 is a realistic or minimal clinically relevant effect size, as median power only exceeded the 80% threshold for this SMD. Realistically, however, effect sizes in psychiatry are more commonly in the range of 0.20-0.60 (Huhn *et al*., 2014). The fact that trials comparing two active treatments (for which the expected effect size is smaller than the expected effect size of an active vs. inactive comparison) were not larger or better powered to detect SMD=0.40 also suggests that trialists do not account for the realistically expected effect size. The apparent tendency to expect very large effects may be, in part, a consequence of biases in the literature, which have led to inflated effect sizes (although we note that our estimated effect sizes also contain this bias). It may also be related to a tradition of calculating power based on the results of small pilot studies, which tend to overestimate effect size (if only statistically significant pilot studies are carried forward into larger studies) (Anderson *et al*., 2017). Effect sizes are not intuitive and commonly-used rules of thumb (e.g. that an SMD of 0.20 is ‘small’, 0.50 is ‘medium’, and 0.80 is ‘large’) may lead researchers to think that fairly large effect sizes (e.g. SMD≥0.50) are more likely than they are or that realistic (but small) effect sizes are clinically irrelevant and not worth powering for. On the other hand, trialists may have sometimes planned an adequate sample size but encountered problems in achieving this (e.g. due to difficulties in recruiting participants within a grant time frame); some of the included trials may also have had low power because they were intended as pilot studies.

Our results are broadly consistent with previous literature. Median power to detect SMD=0.40 was somewhat higher in our study than median power to detect a relative risk of 70% in previous work examining clinical trials in the mental health field (Turner *et al*., 2013), but this may be due to differences in effect size. Our estimate of the median power to detect SMD=0.40 in psychotherapy trials, specifically, is also much lower than previously reported (Flint *et al*., 2015; Sakaluk *et al*., 2019). However, these previous reports estimated either *post hoc* power (which is considered problematic (Gelman, 2019)), or power to detect the meta-analytic effect size for trials comparing psychotherapy to TAU/waitlist. Our estimate for the latter was similar to the median power in these previous studies. Taken together, our findings show that psychotherapy trials are underpowered in general, just like trials of pharmacotherapy and CAM; however, because the effect size of psychotherapy versus TAU/waitlist happens to be large, power to detect this specific effect size is more reasonable than the power to detect the meta-analytic effect size of other intervention-comparator combinations.

Our results also show that statistical power is improving over time, although it remains well below recommended levels. This is in contrast to previous work that found no increase in power over time (Smaldino and McElreath, 2016; Lamberink *et al*., 2018). The difference might be due to the specific set of studies that we examined, suggesting that trends are different in psychiatric clinical trials than in other areas. However, the difference may also be due to methodological differences, such as the fact that we specifically examined continuous efficacy outcomes and looked at power to detect an SMD of 0.40, rather than the ES_MA_.

Consistent with previous work by Turner and colleagues (Turner *et al*., 2013), we also found that underpowered studies tended to yield higher effect sizes. Underpowered studies produce less precise, more variable estimates than adequately-powered studies, but they are as likely to underestimate or to overestimate effect sizes and should not yield a different *average* effect size, all else being equal. This principle is the basis for the funnel plot, which should be symmetrical with greater variability among smaller studies, in the absence of small-study effects (including reporting bias) (Sterne *et al*., 2005). Our finding that underpowered studies yielded higher effect sizes is therefore consistent with reporting bias against underpowered studies with nonsignificant findings. It therefore remains important for meta-analysts to carefully consider the possible biasing effects of including underpowered studies in a meta-analysis and to use methods to mitigate or explore these biasing effects. We note, however, that most meta-analyses in our study included only a few studies, which may make it difficult to address potential problems with underpowered studies.

### Strengths and limitations

An important strength of our study is that we used the highly comprehensive Cochrane dataset. Our analysis was also specific enough to illuminate possible differences among disorders, intervention types, comparators, and outcome types. Because trials are generally only powered to detect their primary outcome, our examination of (continuous) efficacy outcomes separately from safety (and binary efficacy) outcomes make the results more clearly applicable to clinicians. We also examined power from multiple angles, including power to detect both fixed and meta-analytic effect sizes. The fine-grained nature of our analysis adds important new information to previous studies, for instance regarding the differences among comparators.

Our study also has several limitations. First, our analysis was based on the published literature, so estimated effect sizes may be inflated due to reporting bias. Consequently, power to detect the meta-analytic effect size may also have been overestimated, although our sensitivity analysis based on the largest trial did not yield different results. We also used the absolute effect size for comparisons of two active treatments, as the direction of effects is somewhat arbitrary. While this would not affect our estimates of the power to detect the individual ES_MA_, it may have led to an overestimate of ES_avg_ and hence of power to detect the ES_avg_ in our sensitivity analysis. These limitations imply that the problem of low power may actually be even greater than our results already suggest. We also cannot be certain that the direction of effects was fully consistent across all meta-analyses of active treatment vs. controls; if some inconsistency was present, the ES_avg_ and hence power to detect it may have been underestimated. Such inconsistency would also have affected our estimated effect sizes for the different disorders and intervention types (e.g. as plotted in figure 2). Finally, we were unable to determine the actual primary outcome of each included trial.

### Conclusions

In this examination of the comprehensive Cochrane database, we found consistently low power to detect both fixed and meta-analytic effect sizes in trials of interventions for mental disorders. Median power has increased somewhat over time, but remains far below the recommended 80% level. Power was low regardless of the specific disorder or intervention under investigation. Our findings suggest that trialists are implicitly working under the assumption that very large effect sizes are realistic and do not adjust sample sizes for different types of trials, in particular for trials with more versus less active comparators. Importantly, underpowered studies resulted in higher effect sizes than adequately powered studies, consistent with the presence of reporting bias. These findings confirm the urgent need to increase sample sizes in clinical trials and to reduce reporting bias against studies with nonsignificant results to improve the reliability of the published literature.

## Data Availability

All data used in the manuscript are available upon request from the Cochrane Collaboration.

## Acknowledgments

None.

## Notes

Conflicts of Interest: None.

### Competing Interest Statement

The authors have declared no competing interest.

### Funding Statement

This study did not receive any funding.

