## Supplemental tables for "Statistical power in clinical trials of interventions for mood, anxiety, and psychotic disorders"

**Supplemental table 1. Median meta-analysis effect size by disorder and intervention category for continuous efficacy outcomes.**

| Disorder | Intervention | N | | | Meta-analysis SMD |
|---|---|---|---|---|---|
| | | Meta-analyses | Studies | Unique studies | Median (IQR) |
| Overall | Overall | 1993 | 8295 | 2048 | 0.24 (0.09 - 0.46) |
| | CAM | 89 | 312 | 137 | 0.47 (0.19 - 0.69) |
| | PHT | 1259 | 5263 | 1465 | 0.21 (0.08 - 0.41) |
| | PST | 645 | 2720 | 455 | 0.28 (0.11 - 0.53) |
| Anxiety | Overall | 725 | 2895 | 441 | 0.35 (0.15 - 0.58) |
| | CAM | 3 | 11 | 9 | 0.47 (0.04 - 0.67) |
| | PHT | 338 | 1373 | 191 | 0.33 (0.15 - 0.49) |
| | PST | 384 | 1511 | 250 | 0.39 (0.16 - 0.64) |
| Mood | Overall | 558 | 2642 | 855 | 0.19 (0.07 - 0.39) |
| | CAM | 39 | 175 | 66 | 0.41 (0.12 - 0.57) |
| | PHT | 397 | 1724 | 639 | 0.19 (0.07 - 0.39) |
| | PST | 122 | 743 | 150 | 0.15 (0.07 - 0.32) |
| Psychosis | Overall | 710 | 2758 | 754 | 0.17 (0.08 - 0.37) |
| | CAM | 47 | 126 | 62 | 0.54 (0.37 - 0.71) |
| | PHT | 524 | 2166 | 635 | 0.16 (0.07 - 0.35) |
| | PST | 139 | 466 | 57 | 0.15 (0.09 - 0.30) |

*Table notes: For active vs. active comparisons, absolute effect sizes were used, as the experimental and comparator conditions are essentially interchangeable.*

**Supplemental table 2: Median meta-analysis effect size by experimental group and comparator for continuous efficacy outcomes.**

| Experimental group | Comparator | N | | | Meta-analysis SMD |
|---|---|---|---|---|---|
| | | Meta-analyses | Studies | Unique studies | Median (IQR) |
| Pharmacotherapy (mono) | Placebo | 347 | 1490 | 373 | 0.38 (0.23 - 0.56) |
| | Pharmacotherapy (mono) | 641 | 2693 | 970 | 0.14 (0.06 - 0.27) |
| Combination pharmacotherapy | Pharmacotherapy (mono) | 93 | 354 | 127 | 0.34 (0.04 - 0.64) |
| Pharmacotherapy + psychotherapy | Psychotherapy | 183 | 735 | 54 | 0.28 (0.07 - 0.49) |
| | Pharmacotherapy (mono) | 85 | 212 | 21 | 0.35 (0.19 - 0.53) |
| Psychotherapy | TAU/waitlist | 178 | 789 | 193 | 0.58 (0.17 - 0.96) |
| | Psychotherapy | 255 | 920 | 133 | 0.23 (0.11 - 0.39) |
| | Pharmacotherapy | 16 | 64 | 17 | 0.16 (0.11 - 0.22) |
| | Any comparator | 104 | 719 | 108 | 0.17 (0.06 - 0.36) |
| CAM + regular treatment | Pharmacotherapy (mono) | 32 | 82 | 37 | 0.54 (0.42 - 0.74) |
| CAM | Placebo | 15 | 104 | 41 | 0.41 (0.15 - 0.53) |
| | TAU/waitlist | 25 | 68 | 39 | 0.63 (0.31 - 0.91) |
| | Any active comparator | 19 | 65 | 35 | 0.13 (0.08 - 0.37) |

**Supplemental table 3: Median power for a range of effect sizes and the $ES_{MA}$ by disorder and intervention category, for continuous efficacy outcomes**

| Disorder | Inter-vention | Power | | | | |
|---|---|---|---|---|---|---|
| | | SMD=0.20 | SMD=0.40 | SMD=0.60 | SMD=0.80 | $ES_{MA}$ |
| | | Median (IQR) | Median (IQR) | Median (IQR) | Median (IQR) | Median (IQR) |
| Overall | Overall | 0.12 (0.08 - 0.18) | 0.33 (0.19 - 0.54) | 0.63 (0.36 - 0.87) | 0.86 (0.58 - 0.98) | 0.15 (0.07 - 0.44) |
| | CAM | 0.10 (0.08 - 0.14) | 0.26 (0.15 - 0.40) | 0.51 (0.29 - 0.73) | 0.76 (0.47 - 0.93) | 0.27 (0.10 - 0.63) |
| | PHT | 0.12 (0.09 - 0.19) | 0.34 (0.20 - 0.59) | 0.64 (0.40 - 0.91) | 0.87 (0.62 - 0.99) | 0.13 (0.06 - 0.37) |
| | PST | 0.11 (0.08 - 0.15) | 0.28 (0.18 - 0.46) | 0.55 (0.35 - 0.79) | 0.79 (0.56 - 0.96) | 0.19 (0.08 - 0.56) |
| Anxiety | Overall | 0.09 (0.08 - 0.13) | 0.23 (0.17 - 0.39) | 0.46 (0.32 - 0.72) | 0.69 (0.51 - 0.92) | 0.27 (0.10 - 0.62) |
| | CAM | 0.11 (0.10 - 0.14) | 0.31 (0.26 - 0.41) | 0.60 (0.50 - 0.73) | 0.84 (0.74 - 0.91) | 0.31 (0.25 - 0.57) |
| | PHT | 0.09 (0.08 - 0.14) | 0.23 (0.17 - 0.40) | 0.46 (0.32 - 0.73) | 0.69 (0.51 - 0.93) | 0.19 (0.09 - 0.46) |
| | PST | 0.10 (0.08 - 0.13) | 0.24 (0.16 - 0.38) | 0.47 (0.30 - 0.70) | 0.71 (0.48 - 0.91) | 0.37 (0.13 - 0.74) |
| Mood | Overall | 0.12 (0.09 - 0.23) | 0.36 (0.20 - 0.68) | 0.66 (0.39 - 0.95) | 0.89 (0.60 - 1.00) | 0.14 (0.07 - 0.37) |
| | CAM | 0.08 (0.07 - 0.12) | 0.19 (0.13 - 0.33) | 0.36 (0.24 - 0.63) | 0.58 (0.38 - 0.86) | 0.14 (0.07 - 0.28) |
| | PHT | 0.13 (0.09 - 0.22) | 0.36 (0.22 - 0.67) | 0.68 (0.42 - 0.95) | 0.90 (0.65 - 1.00) | 0.16 (0.07 - 0.41) |
| | PST | 0.14 (0.09 - 0.28) | 0.41 (0.20 - 0.79) | 0.73 (0.39 - 0.98) | 0.93 (0.60 - 1.00) | 0.12 (0.06 - 0.31) |
| Psychosis | Overall | 0.12 (0.10 - 0.17) | 0.36 (0.26 - 0.51) | 0.66 (0.50 - 0.84) | 0.89 (0.74 - 0.98) | 0.09 (0.06 - 0.26) |
| | CAM | 0.12 (0.10 - 0.14) | 0.33 (0.26 - 0.42) | 0.63 (0.51 - 0.75) | 0.86 (0.76 - 0.94) | 0.59 (0.33 - 0.97) |
| | PHT | 0.13 (0.10 - 0.19) | 0.39 (0.27 - 0.58) | 0.71 (0.53 - 0.90) | 0.92 (0.77 - 0.99) | 0.09 (0.06 - 0.23) |
| | PST | 0.11 (0.10 - 0.13) | 0.31 (0.24 - 0.38) | 0.59 (0.47 - 0.70) | 0.83 (0.71 - 0.91) | 0.09 (0.07 - 0.15) |

**Supplemental table 4: Power to detect SMD=0.40 and the $ES_{MA}$ for each combination of experimental group and comparator for continuous efficacy outcomes.**

| Experimental group | Comparator | Power | |
|---|---|---|---|
| | | SMD=0.40 | $ES_{MA}$ |
| | | Median (IQR) | Median (IQR) |
| Pharmacotherapy (mono) | Placebo | 0.33 (0.19 - 0.71) | 0.40 (0.15 - 0.79) |
| | Pharmacotherapy (mono) | 0.42 (0.29 - 0.64) | 0.08 (0.06 - 0.17) |
| Combination pharmacotherapy | Pharmacotherapy (mono) | 0.21 (0.14 - 0.33) | 0.19 (0.07 - 0.41) |
| Pharmacotherapy + psychotherapy | Psychotherapy | 0.19 (0.17 - 0.26) | 0.16 (0.07 - 0.30) |
| | Pharmacotherapy (mono) | 0.23 (0.17 - 0.32) | 0.20 (0.09 - 0.39) |
| Psychotherapy | TAU/waitlist | 0.23 (0.16 - 0.34) | 0.66 (0.28 - 0.96) |
| | Psychotherapy | 0.26 (0.17 - 0.36) | 0.10 (0.06 - 0.21) |
| | Pharmacotherapy | 0.22 (0.19 - 0.33) | 0.07 (0.06 - 0.11) |
| | Any comparator | 0.59 (0.31 - 0.84) | 0.20 (0.09 - 0.47) |
| CAM + regular treatment | Pharmacotherapy (mono) | 0.33 (0.23 - 0.52) | 0.60 (0.36 - 0.94) |
| CAM | Placebo | 0.21 (0.13 - 0.31) | 0.15 (0.07 - 0.27) |
| | TAU/waitlist | 0.28 (0.20 - 0.38) | 0.54 (0.25 - 0.96) |
| | Any active comparator | 0.25 (0.15 - 0.48) | 0.10 (0.06 - 0.18) |

**Supplemental table 5: Median meta-analysis effect size by disorder and intervention category for continuous safety outcomes.**

| Disorder | Intervention | N | | | Meta-analysis SMD |
|---|---|---|---|---|---|
| | | Meta-analyses | Studies | Unique studies | Median (IQR) |
| Overall | Overall | 268 | 846 | 242 | 0.13 (0.03 - 0.34) |
| | CAM | 2 | 4 | 4 | 0.14 (-0.06 - 0.35) |
| | PHT | 266 | 842 | 238 | 0.13 (0.03 - 0.34) |
| | PST | 0 | 0 | 0 | - |
| Anxiety | Overall | 3 | 8 | 6 | 0.52 (0.09 - 1.68) |
| | CAM | 0 | 0 | 0 | - |
| | PHT | 3 | 8 | 6 | 0.52 (0.09 - 1.68) |
| | PST | 0 | 0 | 0 | - |
| Mood | Overall | 22 | 49 | 32 | -0.15 (-0.44 - 0.17) |
| | CAM | 0 | 0 | 0 | - |
| | PHT | 22 | 49 | 32 | -0.15 (-0.44 - 0.17) |
| | PST | 0 | 0 | 0 | - |
| Psychosis | Overall | 243 | 789 | 204 | 0.16 (0.05 - 0.35) |
| | CAM | 2 | 4 | 4 | 0.14 (-0.06 - 0.35) |
| | PHT | 241 | 785 | 200 | 0.16 (0.05 - 0.34) |
| | PST | 0 | 0 | 0 | - |

*Table notes: For active vs. active comparisons, absolute effect sizes were used, as the experimental and comparator conditions are essentially interchangeable.*

**Supplemental table 6: Median meta-analysis effect size by disorder and intervention category for binary efficacy outcomes.**

| Disorder | Intervention | N | | | Meta-analysis OR |
|---|---|---|---|---|---|
| | | Meta-analyses | Studies | Unique studies | Median (IQR) |
| Overall | Overall | 1912 | 8804 | 2367 | 1.27 (0.96 - 1.99) |
| | CAM | 38 | 112 | 72 | 1.90 (1.20 - 3.56) |
| | PHT | 1531 | 7168 | 1935 | 1.25 (0.98 - 1.93) |
| | PST | 343 | 1524 | 369 | 1.37 (0.84 - 2.09) |
| Anxiety | Overall | 405 | 1719 | 336 | 0.83 (0.45 - 1.74) |
| | CAM | 1 | 2 | 2 | 12.37 (12.37 - 12.37) |
| | PHT | 214 | 894 | 165 | 0.67 (0.46 - 1.23) |
| | PST | 190 | 823 | 177 | 1.22 (0.44 - 2.10) |
| Mood | Overall | 822 | 3561 | 899 | 1.20 (0.87 - 1.56) |
| | CAM | 21 | 73 | 45 | 1.24 (0.72 - 2.79) |
| | PHT | 724 | 3071 | 767 | 1.19 (0.83 - 1.52) |
| | PST | 77 | 417 | 88 | 1.39 (1.03 - 1.88) |
| Psychosis | Overall | 685 | 3524 | 1132 | 1.68 (1.20 - 2.92) |
| | CAM | 16 | 37 | 25 | 2.86 (1.49 - 6.70) |
| | PHT | 593 | 3203 | 1003 | 1.68 (1.17 - 2.93) |
| | PST | 76 | 284 | 104 | 1.62 (1.27 - 2.37) |

*Table notes: For active vs. active comparisons, we used the inverse for ORs > 1, as the experimental and comparator conditions are essentially interchangeable.*

**Supplemental table 7: Median meta-analysis effect size by disorder and intervention category for binary safety outcomes.**

| Disorder | Intervention | N | | | Meta-analysis SMD |
|---|---|---|---|---|---|
| | | Meta-analyses | Studies | Unique studies | Median (IQR) |
| Overall | **Overall** | 4636 | 18595 | 3250 | 1.36 (1.07 - 1.97) |
| | **CAM** | 107 | 382 | 151 | 1.21 (0.93 - 1.76) |
| | **PHT** | 4390 | 17476 | 2679 | 1.36 (1.07 - 1.98) |
| | **PST** | 139 | 737 | 427 | 1.19 (1.01 - 1.51) |
| Anxiety | **Overall** | 302 | 1263 | 475 | 0.92 (0.51 - 1.49) |
| | **CAM** | 4 | 8 | 4 | 1.11 (0.90 - 1.46) |
| | **PHT** | 216 | 801 | 193 | 0.81 (0.40 - 1.55) |
| | **PST** | 82 | 454 | 285 | 1.13 (0.76 - 1.45) |
| Mood | **Overall** | 1763 | 6491 | 1071 | 1.34 (1.09 - 1.81) |
| | **CAM** | 29 | 138 | 43 | 1.26 (0.95 - 1.37) |
| | **PHT** | 1700 | 6220 | 981 | 1.34 (1.09 - 1.82) |
| | **PST** | 34 | 133 | 47 | 1.21 (1.17 - 1.71) |
| Psychosis | **Overall** | 2571 | 10841 | 1704 | 1.40 (1.09 - 2.11) |
| | **CAM** | 74 | 236 | 104 | 1.20 (0.93 - 2.19) |
| | **PHT** | 2474 | 10455 | 1505 | 1.41 (1.09 - 2.10) |
| | **PST** | 23 | 150 | 95 | 1.38 (1.17 - 2.42) |

*Table notes: For active vs. active comparisons, we used the inverse for ORs > 1, as the experimental and comparator conditions are essentially interchangeable.*

**Supplemental table 8: Median meta-analysis effect size by experimental group and comparator for continuous safety outcomes.**

| Experimental group | Comparator | N | | | Meta-analysis SMD |
|---|---|---|---|---|---|
| | | Meta-analyses | Studies | Unique studies | Median (IQR) |
| Pharmacotherapy (mono) | Placebo | 42 | 150 | 36 | -0.14 (-0.40 - -0.02) |
| | Pharmacotherapy (mono) | 218 | 677 | 197 | 0.21 (0.08 - 0.43) |
| Combination pharmacotherapy | Pharmacotherapy (mono) | 6 | 15 | 13 | -0.38 (-1.32 - -0.16) |
| CAM + regular treatment | Pharmacotherapy (mono) | 2 | 4 | 4 | 0.14 (-0.06 - 0.35) |

**Supplemental table 9: Median meta-analysis effect size by experimental group and comparator for binary efficacy outcomes.**

| Experimental group | Comparator | N | | | Meta-analysis OR |
|---|---|---|---|---|---|
| | | Meta-analyses | Studies | Unique studies | Median (IQR) |
| Pharmacotherapy (mono) | Placebo | 540 | 3017 | 593 | 1.16 (0.46 - 3.09) |
| | Pharmacotherapy (mono) | 771 | 3090 | 1244 | 1.29 (1.12 - 1.62) |
| Combination pharmacotherapy | Pharmacotherapy (mono) | 119 | 622 | 120 | 1.46 (0.62 - 2.10) |
| Pharmacotherapy + psychotherapy | Psychotherapy | 104 | 445 | 42 | 0.67 (0.60 - 0.96) |
| | Pharmacotherapy (mono) | 54 | 134 | 17 | 0.57 (0.42 - 0.80) |
| Psychotherapy | TAU/waitlist | 92 | 479 | 183 | 1.35 (0.25 - 2.32) |
| | Psychotherapy | 110 | 471 | 122 | 1.60 (1.24 - 2.32) |
| | Pharmacotherapy | 46 | 242 | 22 | 1.58 (1.22 - 1.89) |
| | Any comparator | 37 | 190 | 45 | 1.71 (1.27 - 2.17) |
| CAM + regular treatment | Pharmacotherapy (mono) | 11 | 26 | 19 | 2.77 (1.75 - 5.86) |
| CAM | Placebo/TAU/waitlist | 8 | 33 | 25 | 0.69 (0.49 - 2.34) |
| | Any active comparator | 20 | 55 | 35 | 1.80 (1.22 - 3.16) |

**Supplemental table 10: Median meta-analysis effect size by experimental group and comparator for binary safety outcomes.**

| Experimental group | Comparator | N | | | Meta-analysis OR |
|---|---|---|---|---|---|
| | | Meta-analyses | Studies | Unique studies | Median (IQR) |
| Pharmacotherapy (mono) | Placebo | 909 | 3721 | 675 | 0.81 (0.45 - 1.28) |
| | Pharmacotherapy (mono) | 3266 | 12780 | 1875 | 1.49 (1.20 - 2.20) |
| Combination pharmacotherapy | Pharmacotherapy (mono) | 152 | 700 | 184 | 0.83 (0.47 - 1.31) |
| Pharmacotherapy + psychotherapy | Psychotherapy | 65 | 278 | 49 | 0.78 (0.32 - 0.92) |
| | Pharmacotherapy (mono) | 28 | 78 | 16 | 1.23 (0.95 - 1.52) |
| Psychotherapy | TAU/waitlist | 38 | 297 | 241 | 0.94 (0.63 - 1.25) |
| | Psychotherapy | 48 | 260 | 149 | 1.28 (1.14 - 1.74) |
| | Pharmacotherapy | 12 | 51 | 18 | 1.55 (1.18 - 1.98) |
| | Any comparator | 9 | 41 | 31 | 1.16 (0.74 - 1.30) |
| CAM + regular treatment | TAU/waitlist | 3 | 12 | 10 | 1.23 (1.05 - 1.82) |
| | Pharmacotherapy (mono) | 51 | 154 | 48 | 1.18 (0.93 - 2.52) |
| CAM | Placebo/TAU/waitlist | 31 | 161 | 76 | 1.05 (0.64 - 1.24) |
| | Any active comparator | 24 | 62 | 30 | 1.43 (1.26 - 2.09) |

**Supplemental table 11: Median power for a range of effect sizes and the $ES_{MA}$ by disorder and intervention category, for continuous safety outcomes**

| Disorder | Inter-vention | Power | | | | |
|---|---|---|---|---|---|---|
| | | SMD=0.20 | SMD=0.40 | SMD=0.60 | SMD=0.80 | $ES_{MA}$ |
| | | Median (IQR) | Median (IQR) | Median (IQR) | Median (IQR) | Median (IQR) |
| Overall | Overall | 0.28 (0.13 - 0.45) | 0.79 (0.39 - 0.95) | 0.99 (0.71 - 1.00) | 1.00 (0.92 - 1.00) | 0.20 (0.08 - 0.79) |
| | CAM | 0.12 (0.09 - 0.17) | 0.33 (0.23 - 0.49) | 0.61 (0.46 - 0.81) | 0.82 (0.69 - 0.96) | 0.30 (0.20 - 0.52) |
| | PHT | 0.29 (0.13 - 0.45) | 0.80 (0.39 - 0.95) | 0.99 (0.71 - 1.00) | 1.00 (0.92 - 1.00) | 0.20 (0.08 - 0.79) |
| | PST | - | - | - | - | - |
| Anxiety | Overall | 0.54 (0.52 - 0.55) | 0.98 (0.98 - 0.99) | 1.00 (1.00 - 1.00) | 1.00 (1.00 - 1.00) | 1.00 (0.94 - 1.00) |
| | CAM | - | - | - | - | - |
| | PHT | 0.54 (0.52 - 0.55) | 0.98 (0.98 - 0.99) | 1.00 (1.00 - 1.00) | 1.00 (1.00 - 1.00) | 1.00 (0.94 - 1.00) |
| | PST | - | - | - | - | - |
| Mood | Overall | 0.34 (0.20 - 0.39) | 0.87 (0.61 - 0.92) | 1.00 (0.92 - 1.00) | 1.00 (0.99 - 1.00) | 0.76 (0.18 - 0.99) |
| | CAM | - | - | - | - | - |
| | PHT | 0.34 (0.20 - 0.39) | 0.87 (0.61 - 0.92) | 1.00 (0.92 - 1.00) | 1.00 (0.99 - 1.00) | 0.76 (0.18 - 0.99) |
| | PST | - | - | - | - | - |
| Psychosis | Overall | 0.28 (0.12 - 0.45) | 0.78 (0.35 - 0.95) | 0.98 (0.66 - 1.00) | 1.00 (0.88 - 1.00) | 0.19 (0.08 - 0.73) |
| | CAM | 0.12 (0.09 - 0.17) | 0.33 (0.23 - 0.49) | 0.61 (0.46 - 0.81) | 0.82 (0.69 - 0.96) | 0.30 (0.20 - 0.52) |
| | PHT | 0.28 (0.12 - 0.45) | 0.79 (0.35 - 0.95) | 0.99 (0.66 - 1.00) | 1.00 (0.88 - 1.00) | 0.19 (0.08 - 0.73) |
| | PST | - | - | - | - | - |

**Supplemental table 12: Median power for a range of effect sizes and the $ES_{MA}$ by disorder and intervention category, for binary efficacy outcomes**

| Disorder | Inter-vention | Power | | | | |
|---|---|---|---|---|---|---|
| | | OR=1.5 | OR=2.0 | OR=3.0 | OR=4.5 | $ES_{MA}$ |
| | | Median (IQR) | Median (IQR) | Median (IQR) | Median (IQR) | Median (IQR) |
| Overall | Overall | 0.11 (0.08 - 0.17) | 0.22 (0.15 - 0.40) | 0.47 (0.30 - 0.77) | 0.72 (0.49 - 0.95) | 0.15 (0.07 - 0.38) |
| | CAM | 0.10 (0.08 - 0.14) | 0.20 (0.12 - 0.33) | 0.42 (0.23 - 0.66) | 0.64 (0.36 - 0.90) | 0.15 (0.08 - 0.36) |
| | PHT | 0.11 (0.08 - 0.18) | 0.23 (0.15 - 0.43) | 0.48 (0.31 - 0.80) | 0.73 (0.50 - 0.96) | 0.15 (0.06 - 0.39) |
| | PST | 0.10 (0.08 - 0.14) | 0.21 (0.15 - 0.31) | 0.43 (0.29 - 0.64) | 0.66 (0.47 - 0.88) | 0.15 (0.07 - 0.36) |
| Anxiety | Overall | 0.11 (0.09 - 0.15) | 0.21 (0.16 - 0.34) | 0.44 (0.32 - 0.67) | 0.66 (0.51 - 0.90) | 0.20 (0.09 - 0.52) |
| | CAM | 0.06 (0.06 - 0.06) | 0.07 (0.07 - 0.07) | 0.10 (0.10 - 0.10) | 0.13 (0.13 - 0.14) | 0.20 (0.20 - 0.21) |
| | PHT | 0.11 (0.09 - 0.15) | 0.23 (0.16 - 0.35) | 0.48 (0.32 - 0.70) | 0.72 (0.52 - 0.91) | 0.17 (0.10 - 0.55) |
| | PST | 0.10 (0.09 - 0.14) | 0.21 (0.16 - 0.32) | 0.43 (0.32 - 0.65) | 0.65 (0.50 - 0.88) | 0.23 (0.09 - 0.49) |
| Mood | Overall | 0.12 (0.09 - 0.22) | 0.26 (0.16 - 0.52) | 0.54 (0.33 - 0.88) | 0.78 (0.53 - 0.99) | 0.13 (0.07 - 0.32) |
| | CAM | 0.10 (0.07 - 0.15) | 0.19 (0.12 - 0.34) | 0.40 (0.22 - 0.67) | 0.62 (0.34 - 0.90) | 0.13 (0.08 - 0.29) |
| | PHT | 0.13 (0.09 - 0.24) | 0.29 (0.17 - 0.57) | 0.59 (0.35 - 0.91) | 0.83 (0.57 - 0.99) | 0.15 (0.07 - 0.35) |
| | PST | 0.10 (0.07 - 0.13) | 0.19 (0.11 - 0.29) | 0.39 (0.21 - 0.60) | 0.61 (0.36 - 0.85) | 0.09 (0.06 - 0.17) |
| Psychosis | Overall | 0.10 (0.08 - 0.14) | 0.20 (0.14 - 0.32) | 0.43 (0.28 - 0.67) | 0.69 (0.46 - 0.91) | 0.14 (0.06 - 0.39) |
| | CAM | 0.11 (0.09 - 0.13) | 0.24 (0.17 - 0.31) | 0.53 (0.36 - 0.65) | 0.81 (0.60 - 0.91) | 0.33 (0.09 - 0.52) |
| | PHT | 0.10 (0.08 - 0.14) | 0.19 (0.14 - 0.32) | 0.42 (0.27 - 0.67) | 0.68 (0.45 - 0.91) | 0.14 (0.06 - 0.40) |
| | PST | 0.11 (0.09 - 0.13) | 0.23 (0.17 - 0.30) | 0.49 (0.35 - 0.64) | 0.74 (0.56 - 0.89) | 0.14 (0.07 - 0.30) |

**Supplemental table 13: Median power for a range of effect sizes and the $ES_{MA}$ by disorder and intervention category, for binary safety outcomes**

| Disorder | Inter-vention | Power | | | | | |
|---|---|---|---|---|---|---|---|
| | | OR=1.5 | OR=2.0 | OR=3.0 | OR=4.5 | $ES_{MA}$ | $ES_{avg}$ |
| | | Median (IQR) | Median (IQR) | Median (IQR) | Median (IQR) | Median (IQR) | Median (IQR) |
| Overall | Overall | 0.10 (0.07 - 0.15) | 0.21 (0.13 - 0.38) | 0.47 (0.27 - 0.79) | 0.76 (0.49 - 0.98) | 0.08 (0.06 - 0.20) | 0.09 (0.06 - 0.13) |
| | CAM | 0.07 (0.06 - 0.09) | 0.11 (0.08 - 0.18) | 0.24 (0.15 - 0.39) | 0.43 (0.27 - 0.66) | 0.06 (0.05 - 0.09) | 0.05 (0.05 - 0.06) |
| | PHT | 0.10 (0.07 - 0.16) | 0.21 (0.13 - 0.40) | 0.48 (0.28 - 0.81) | 0.77 (0.50 - 0.98) | 0.08 (0.06 - 0.21) | 0.09 (0.07 - 0.14) |
| | PST | 0.09 (0.07 - 0.12) | 0.17 (0.12 - 0.26) | 0.38 (0.24 - 0.58) | 0.63 (0.42 - 0.87) | 0.07 (0.05 - 0.09) | 0.05 (0.05 - 0.06) |
| Anxiety | Overall | 0.09 (0.07 - 0.13) | 0.18 (0.12 - 0.31) | 0.41 (0.26 - 0.67) | 0.69 (0.45 - 0.92) | 0.08 (0.06 - 0.22) | 0.06 (0.05 - 0.08) |
| | CAM | 0.07 (0.07 - 0.09) | 0.13 (0.11 - 0.20) | 0.31 (0.22 - 0.47) | 0.58 (0.40 - 0.78) | 0.08 (0.05 - 0.11) | 0.07 (0.07 - 0.09) |
| | PHT | 0.09 (0.07 - 0.14) | 0.19 (0.12 - 0.33) | 0.43 (0.26 - 0.70) | 0.72 (0.46 - 0.94) | 0.12 (0.06 - 0.43) | 0.07 (0.06 - 0.10) |
| | PST | 0.09 (0.07 - 0.12) | 0.18 (0.12 - 0.28) | 0.39 (0.26 - 0.62) | 0.65 (0.44 - 0.88) | 0.07 (0.05 - 0.10) | 0.05 (0.05 - 0.06) |
| Mood | Overall | 0.11 (0.08 - 0.18) | 0.26 (0.15 - 0.46) | 0.60 (0.35 - 0.87) | 0.88 (0.60 - 0.99) | 0.09 (0.06 - 0.19) | 0.10 (0.07 - 0.16) |
| | CAM | 0.08 (0.07 - 0.10) | 0.13 (0.10 - 0.20) | 0.26 (0.18 - 0.40) | 0.45 (0.31 - 0.64) | 0.06 (0.05 - 0.08) | 0.05 (0.05 - 0.06) |
| | PHT | 0.12 (0.08 - 0.19) | 0.27 (0.16 - 0.47) | 0.62 (0.36 - 0.88) | 0.90 (0.63 - 0.99) | 0.09 (0.06 - 0.20) | 0.10 (0.07 - 0.16) |
| | PST | 0.07 (0.06 - 0.09) | 0.12 (0.08 - 0.18) | 0.22 (0.13 - 0.41) | 0.41 (0.23 - 0.70) | 0.06 (0.05 - 0.09) | 0.05 (0.05 - 0.06) |
| Psychosis | Overall | 0.09 (0.07 - 0.14) | 0.18 (0.12 - 0.32) | 0.41 (0.24 - 0.70) | 0.69 (0.44 - 0.94) | 0.08 (0.06 - 0.20) | 0.08 (0.06 - 0.12) |
| | CAM | 0.07 (0.06 - 0.09) | 0.10 (0.08 - 0.17) | 0.21 (0.14 - 0.39) | 0.41 (0.24 - 0.67) | 0.05 (0.05 - 0.10) | 0.06 (0.05 - 0.07) |
| | PHT | 0.09 (0.07 - 0.14) | 0.19 (0.12 - 0.33) | 0.42 (0.25 - 0.71) | 0.70 (0.45 - 0.95) | 0.08 (0.06 - 0.21) | 0.09 (0.06 - 0.12) |
| | PST | 0.10 (0.08 - 0.12) | 0.20 (0.15 - 0.28) | 0.45 (0.33 - 0.63) | 0.74 (0.58 - 0.90) | 0.06 (0.05 - 0.08) | 0.05 (0.05 - 0.06) |

**Supplemental table 14: Power for each combination of experimental group and comparator for continuous safety outcomes.**

| Experimental group | Comparator | Power | |
|---|---|---|---|
| | | SMD=0.40 | $ES_{MA}$ |
| | | Median (IQR) | Median (IQR) |
| Pharmacotherapy (mono) | Placebo | 0.85 (0.72 - 0.95) | 0.16 (0.07 - 0.93) |
| | Pharmacotherapy (mono) | 0.75 (0.28 - 0.95) | 0.21 (0.09 - 0.74) |
| Combination pharmacotherapy | Pharmacotherapy (mono) | 0.91 (0.40 - 0.96) | 0.77 (0.20 - 1.00) |
| Pharmacotherapy + psychotherapy | Psychotherapy | - | - |
| | Pharmacotherapy (mono) | - | - |
| Psychotherapy | TAU/waitlist | - | - |
| | Psychotherapy | - | - |
| | Pharmacotherapy | - | - |
| | Any comparator | - | - |
| CAM + regular treatment | TAU/waitlist | - | - |
| | Pharmacotherapy (mono) | 0.33 (0.23 - 0.49) | 0.30 (0.20 - 0.52) |
| CAM | Placebo/TAU/waitlist | - | - |
| | Any active comparator | - | - |

**Supplemental table 15: Power for each combination of experimental group and comparator for binary efficacy outcomes.**

| Experimental group | Comparator | Power | |
|---|---|---|---|
| | | SMD=0.40 | $ES_{MA}$ |
| | | Median (IQR) | Median (IQR) |
| Pharmacotherapy (mono) | Placebo | 0.22 (0.13 - 0.45) | 0.43 (0.19 - 0.79) |
| | Pharmacotherapy (mono) | 0.25 (0.17 - 0.53) | 0.07 (0.05 - 0.13) |
| Combination pharmacotherapy | Pharmacotherapy (mono) | 0.21 (0.14 - 0.30) | 0.19 (0.10 - 0.33) |
| Pharmacotherapy + psychotherapy | Psychotherapy | 0.20 (0.16 - 0.31) | 0.12 (0.08 - 0.17) |
| | Pharmacotherapy (mono) | 0.24 (0.15 - 0.39) | 0.23 (0.11 - 0.36) |
| Psychotherapy | TAU/waitlist | 0.22 (0.16 - 0.36) | 0.51 (0.26 - 0.84) |
| | Psychotherapy | 0.17 (0.12 - 0.25) | 0.08 (0.06 - 0.18) |
| | Pharmacotherapy | 0.18 (0.16 - 0.24) | 0.09 (0.07 - 0.15) |
| | Any comparator | 0.40 (0.23 - 0.57) | 0.15 (0.08 - 0.35) |
| CAM + regular treatment | TAU/waitlist | - | - |
| | Pharmacotherapy (mono) | 0.25 (0.17 - 0.30) | 0.53 (0.11 - 0.87) |
| CAM | Placebo/TAU/waitlist | 0.19 (0.11 - 0.28) | 0.08 (0.07 - 0.19) |
| | Any active comparator | 0.20 (0.12 - 0.37) | 0.15 (0.07 - 0.35) |

**Supplemental table 16: Power for each combination of experimental group and comparator for binary safety outcomes.**

| Experimental group | Comparator | Power | |
|---|---|---|---|
| | | SMD=0.40 | $ES_{MA}$ |
| | | Median (IQR) | Median (IQR) |
| Pharmacotherapy (mono) | Placebo | 0.22 (0.13 - 0.44) | 0.11 (0.06 - 0.32) |
| | Pharmacotherapy (mono) | 0.21 (0.13 - 0.40) | 0.09 (0.06 - 0.25) |
| Combination pharmacotherapy | Pharmacotherapy (mono) | 0.14 (0.10 - 0.25) | 0.06 (0.05 - 0.11) |
| Pharmacotherapy + psychotherapy | Psychotherapy | 0.16 (0.13 - 0.23) | 0.08 (0.06 - 0.16) |
| | Pharmacotherapy (mono) | 0.19 (0.14 - 0.29) | 0.07 (0.06 - 0.10) |
| Psychotherapy | TAU/waitlist | 0.17 (0.12 - 0.26) | 0.07 (0.06 - 0.10) |
| | Psychotherapy | 0.15 (0.10 - 0.22) | 0.06 (0.05 - 0.07) |
| | Pharmacotherapy | 0.19 (0.15 - 0.29) | 0.07 (0.06 - 0.11) |
| | Any comparator | 0.28 (0.16 - 0.49) | 0.06 (0.05 - 0.08) |
| CAM + regular treatment | TAU/waitlist | 0.07 (0.06 - 0.08) | 0.05 (0.05 - 0.05) |
| | Pharmacotherapy (mono) | 0.10 (0.08 - 0.16) | 0.05 (0.05 - 0.12) |
| CAM | Placebo/TAU/waitlist | 0.14 (0.09 - 0.20) | 0.06 (0.05 - 0.07) |
| | Any active comparator | 0.12 (0.09 - 0.17) | 0.06 (0.05 - 0.15) |

**Supplemental table 17: $ES_{avg}$ for each intervention-comparator combination, for continuous efficacy outcomes only (sensitivity analyses)**

| Experimental group | Comparator | Number of meta-analyses | $ES_{avg}$ |
|---|---|---|---|
| Pharmacotherapy (mono) | Placebo | 347 | 0.34 |
| | Pharmacotherapy (mono) | 641 | 0.14 |
| Combination pharmacotherapy | Pharmacotherapy (mono) | 93 | 0.30 |
| Pharmacotherapy + psychotherapy | Psychotherapy | 183 | 0.29 |
| | Pharmacotherapy (mono) | 85 | 0.28 |
| Psychotherapy | TAU/waitlist | 178 | 0.56 |
| | Psychotherapy | 255 | 0.23 |
| | Pharmacotherapy | 16 | 0.18 |
| | Any comparator | 104 | 0.17 |
| CAM + regular treatment | Pharmacotherapy (mono) | 32 | 0.59 |
| CAM | Placebo | 15 | 0.25 |
| | TAU/waitlist | 25 | 0.49 |
| | Any active comparator | 19 | 0.16 |

**Supplemental table 18: Median power by disorder and intervention category in sensitivity analyses, for continuous efficacy outcomes (sensitivity analyses)**

| Disorder | Inter-vention | Power | | |
|---|---|---|---|---|
| | | $ES_{avg}$ | $ES_{MA}$ (meta-analyses with $ES_{MA}$<0.20 excluded) | $ES_{MA}$ ($ES_{MA}$ based on the largest RCT in each meta-analysis) |
| | | Median (IQR) | Median (IQR) | Median (IQR) |
| Overall | Overall | 0.14 (0.09 - 0.25) | 0.36 (0.18 - 0.71) | 0.13 (0.06 - 0.40) |
| | CAM | 0.24 (0.09 - 0.51) | 0.40 (0.20 - 0.81) | 0.15 (0.06 - 0.74) |
| | PHT | 0.12 (0.09 - 0.21) | 0.33 (0.17 - 0.68) | 0.13 (0.06 - 0.39) |
| | PST | 0.16 (0.11 - 0.31) | 0.41 (0.19 - 0.76) | 0.12 (0.07 - 0.41) |
| Anxiety | Overall | 0.16 (0.11 - 0.32) | 0.39 (0.19 - 0.74) | 0.24 (0.10 - 0.59) |
| | CAM | 0.15 (0.13 - 0.58) | 0.31 (0.25 - 0.57) | 0.26 (0.21 - 0.53) |
| | PHT | 0.14 (0.11 - 0.25) | 0.30 (0.16 - 0.59) | 0.26 (0.12 - 0.55) |
| | PST | 0.18 (0.11 - 0.39) | 0.51 (0.24 - 0.86) | 0.21 (0.09 - 0.69) |
| Mood | Overall | 0.15 (0.09 - 0.26) | 0.33 (0.17 - 0.64) | 0.10 (0.06 - 0.28) |
| | CAM | 0.10 (0.07 - 0.19) | 0.23 (0.14 - 0.48) | 0.07 (0.06 - 0.18) |
| | PHT | 0.16 (0.10 - 0.27) | 0.35 (0.19 - 0.68) | 0.12 (0.07 - 0.32) |
| | PST | 0.14 (0.09 - 0.25) | 0.31 (0.16 - 0.58) | 0.07 (0.05 - 0.20) |
| Psychosis | Overall | 0.11 (0.08 - 0.17) | 0.34 (0.17 - 0.75) | 0.09 (0.06 - 0.29) |
| | CAM | 0.53 (0.39 - 0.69) | 0.63 (0.35 - 0.99) | 0.75 (0.16 - 0.97) |
| | PHT | 0.10 (0.08 - 0.14) | 0.36 (0.18 - 0.78) | 0.09 (0.05 - 0.28) |
| | PST | 0.15 (0.12 - 0.17) | 0.17 (0.12 - 0.38) | 0.09 (0.06 - 0.22) |

*Table notes: Several sensitivity analyses were performed. First, we calculated power to detect the $ES_{avg}$, the average meta-analytic effect size for each experimental group – comparator combination. Secondly, we calculated power to detect the $ES_{MA}$ after exclusion of meta-analyses with an* *absolute effect size smaller than 0.20. Thirdly, we calculated power to detect the $ES_{MA}$ based on the largest RCT in each meta-analysis, to reduce the influence of publication bias on the $ES_{MA}$.*

**Supplemental table 19: Sensitivity analysis of median power for a range of effect sizes and the $ES_{MA}$ by disorder and intervention category, for continuous efficacy outcomes, only including each study once.**

| Disorder | Inter-vention | Power | | | | |
|---|---|---|---|---|---|---|
| | | SMD=0.20 | SMD=0.40 | SMD=0.60 | SMD=0.80 | $ES_{MA}$ |
| | | Median (IQR) | Median (IQR) | Median (IQR) | Median (IQR) | Median (IQR) |
| Overall | Overall | 0.12 (0.09 - 0.19) | 0.33 (0.21 - 0.59) | 0.63 (0.40 - 0.91) | 0.86 (0.62 - 0.99) | 0.13 (0.06 - 0.46) |
| | CAM | 0.11 (0.08 - 0.14) | 0.28 (0.17 - 0.41) | 0.55 (0.33 - 0.73) | 0.79 (0.53 - 0.93) | 0.31 (0.10 - 0.67) |
| | PHT | 0.13 (0.09 - 0.23) | 0.37 (0.23 - 0.68) | 0.68 (0.45 - 0.95) | 0.90 (0.68 - 1.00) | 0.10 (0.06 - 0.31) |
| | PST | 0.10 (0.08 - 0.15) | 0.27 (0.17 - 0.46) | 0.53 (0.33 - 0.80) | 0.77 (0.53 - 0.96) | 0.30 (0.10 - 0.77) |
| Anxiety | Overall | 0.10 (0.08 - 0.15) | 0.25 (0.17 - 0.44) | 0.49 (0.31 - 0.77) | 0.74 (0.50 - 0.95) | 0.34 (0.12 - 0.83) |
| | CAM | 0.11 (0.09 - 0.12) | 0.31 (0.23 - 0.32) | 0.59 (0.46 - 0.61) | 0.83 (0.69 - 0.85) | 0.30 (0.23 - 0.40) |
| | PHT | 0.10 (0.08 - 0.18) | 0.26 (0.17 - 0.54) | 0.51 (0.32 - 0.87) | 0.75 (0.51 - 0.99) | 0.24 (0.10 - 0.61) |
| | PST | 0.10 (0.08 - 0.13) | 0.24 (0.16 - 0.36) | 0.47 (0.30 - 0.68) | 0.71 (0.48 - 0.90) | 0.56 (0.15 - 0.92) |
| Mood | Overall | 0.14 (0.09 - 0.27) | 0.40 (0.21 - 0.77) | 0.73 (0.41 - 0.98) | 0.93 (0.63 - 1.00) | 0.12 (0.07 - 0.33) |
| | CAM | 0.09 (0.07 - 0.12) | 0.21 (0.14 - 0.34) | 0.40 (0.25 - 0.64) | 0.63 (0.40 - 0.87) | 0.14 (0.07 - 0.27) |
| | PHT | 0.15 (0.10 - 0.29) | 0.45 (0.24 - 0.80) | 0.79 (0.47 - 0.99) | 0.96 (0.70 - 1.00) | 0.11 (0.06 - 0.31) |
| | PST | 0.12 (0.08 - 0.25) | 0.34 (0.18 - 0.74) | 0.64 (0.34 - 0.97) | 0.87 (0.54 - 1.00) | 0.16 (0.07 - 0.36) |
| Psychosis | Overall | 0.12 (0.09 - 0.17) | 0.34 (0.24 - 0.50) | 0.64 (0.46 - 0.84) | 0.87 (0.70 - 0.97) | 0.09 (0.06 - 0.38) |
| | CAM | 0.12 (0.10 - 0.14) | 0.33 (0.24 - 0.42) | 0.63 (0.47 - 0.75) | 0.86 (0.71 - 0.94) | 0.63 (0.34 - 0.98) |
| | PHT | 0.12 (0.10 - 0.17) | 0.35 (0.24 - 0.52) | 0.66 (0.46 - 0.85) | 0.88 (0.70 - 0.98) | 0.08 (0.06 - 0.24) |
| | PST | 0.11 (0.09 - 0.14) | 0.28 (0.23 - 0.40) | 0.55 (0.45 - 0.72) | 0.79 (0.69 - 0.92) | 0.15 (0.08 - 0.72) |

**Supplemental figure 1: Median power by year of trial publication (in 5-year intervals)**

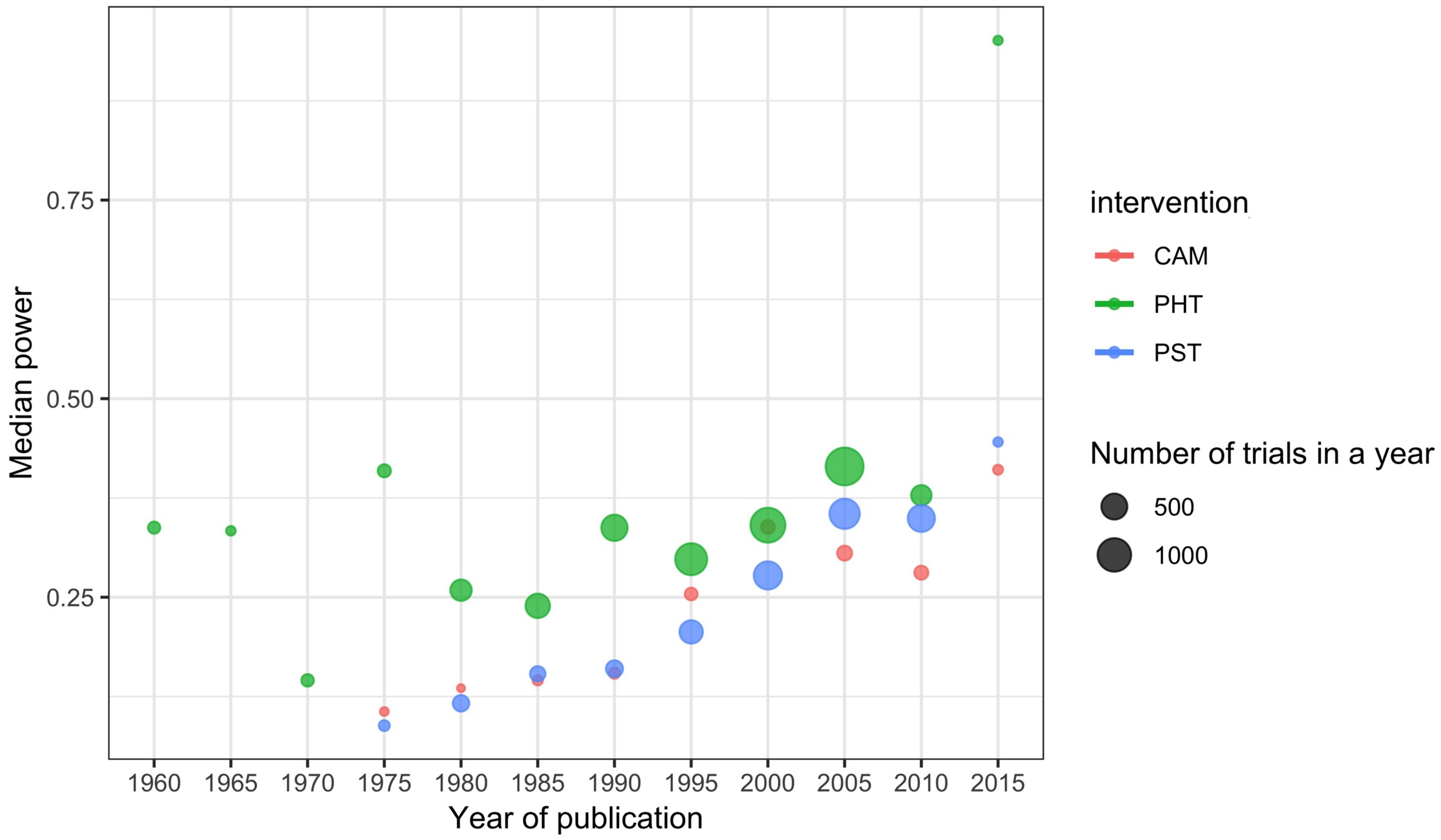
